# Analytic Challenges in Clinical Trials of Early Alzheimer’s Disease

**DOI:** 10.1101/2024.05.23.24307810

**Authors:** Craig Mallinckrodt, Ilya Lipkovich, Suzanne Hendrix, Sam Dickson, Geert Molenberghs

## Abstract

The present investigation assessed how the heavily right-skewed data seen in recently reported results in Alzheimer’s Disease (AD) clinical trials influenced treatment contrasts when data were analyzed via the typical mixed-effects model for repeated measures (MMRM) versus robust regression (RR) and the non-parametric Hodges-Lehman estimator (HL).

Results in simulated data patterned after AD trials showed that imbalance across treatment arms in the number of patients in the extreme right tail (those with rapid disease progression) frequently occurred by chance alone. Each analysis method controlled Type I error at or below the nominal level. The RR analysis yielded smaller standard errors, and more power than MMRM and HL. In datasets with appreciable imbalance in the number of rapid progressing patients, MMRM results favored the treatment arm with fewer rapid progressors. Results from HL showed the same trend, but to a lesser degree. Robust regression yielded similar results regardless of the ratio of rapid progressors. Although more research is needed over a wider range of scenarios, it should not be assumed that MMRM is the optimal approach for trials in early Alzheimer’s Disease.

## 1. Introduction

Alzheimer’s Disease (AD) is a complex and heterogeneous neurodegenerative disease. Developing novel treatments for AD requires accurate diagnosis of the disease, accurate measurement of disease progression, and reliable analysis of the data. An important analytic challenge has emerged in recent AD clinical trials. Heavily skewed data with small imbalances in the number of rapidly progressing patients has had a relatively large impact on differences between treatments in mean change to endpoint in two recently completed, identically designed trials in AD ^1, 3^ p^72, 4^. Results from the primary and multiplicity adjusted secondary outcomes of these studies are provided in the appendix (Table A1)^1^.

The primary analysis in these studies was MMRM, and the primary estimand was based on the treatment policy strategy for dealing with the intercurrent event of early study drug discontinuation^1^. Both the sponsor and FDA conducted post-hoc sensitivity analyses utilizing transformations, non-parametric methods, and robust regression to deal with the unexpectedly large departure from normality, a key assumption in ANCOVA, MMRM, and similar analyses ^1, 2 p 340, 3^.

Rapid progressing patients do not differ from other patients in demographic or baseline disease characteristics, comorbidities, concomitant medications, or the incidence of adverse events ^3,4^. There is no single clinical feature that differentiates rapidly progressing patients from other patients ^5^. Therefore, rapid progressing patients (and the resultant skewed data) are part of the reality of Alzheimer’s Disease and after the fact it is too late to address them in a completed randomized trial ^2 p340^ . However, current practice does not often include assessments of and sensitivity analyses for rapid progressors / outliers. Therefore, the analytic challenge is how to plan for rapid progressing patients in the analyses of Alzheimer’s clinical trial data.

The primary purpose of the present investigation is to compare MMRM with a non-parametric and a robust regression approach in data with and without rapid progressing patients to provide insight on ways to proactively deal with rapid progressing patients and the highly skewed data that has been encountered in AD clinical trials. The remainder of this paper is organized as follows: Section 2 provides an overview of the consequences of and analytic approaches for dealing with skewed data; Section 3 outlines the design and analysis of a simulation study to address the objective of comparing MMRM with a non-parametric and robust regression analytic approaches; Section 4 details results of the simulation study; and, Section 5 discusses those results in the context of the strengths and limitations of the simulation study.

## 2. Consequences of and analytic approaches for dealing with skewed data

The statistical consequence of rapidly progressing patients is outliers / non-normality of the data^1, 2, pp 69-72, 3^. Figure 1 shows the distribution of changes from baseline to week 78 in the primary outcome (CDR-sb) of the two identically designed clinical trials noted in Section 1^4^. The data were heavily right skewed, with ∼1% of patients having > 8-point change from baseline. The distribution of other outcomes was similarly skewed ^1, 2^.

**Figure 1.**
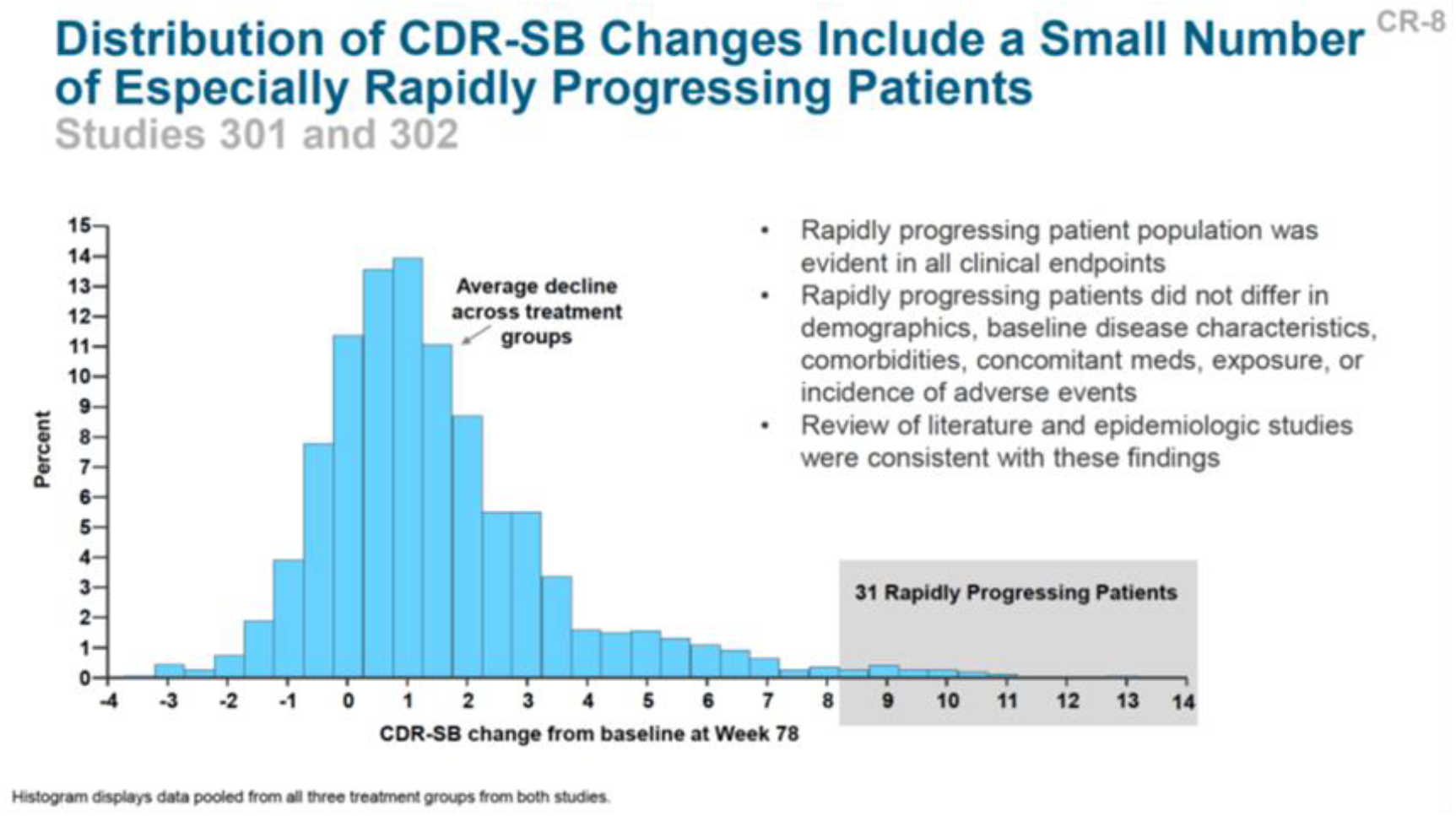
Distribution of the primary outcome, CDR-sb in the aducanumab clinical trials^4^

Statistical theory suggests methods other than MMRM may be useful to consider when data are heavily skewed ^6^. Via the central limit theorem, in large trials the concern regarding non-normal data is not bias, the concern is stability of results ^7, 8, 9, 10^. However, in smaller trials bias may also be a concern.

The potential influence of heavily skewed data (or outliers) can be put into perspective by noting that an outlier with 3-fold the error magnitude of a typical observation contributes 9-fold (3^2^) times as much to the squared error loss, and an outlier with 5-fold the error magnitude contributes 25-fold. Therefore, even a few outliers can increase variance substantially. Maximum likelihood methods such as MMRM are robust to departures from normality in the sense that the Type I error rate does not increase under violations of normality so long as sample sizes are not small (∼40 patients per arm or larger). However, estimates of individual parameters and Type II error may not be so robust.

General categories of methodology for dealing with skewed distributions include robust methods and non-parametric methods. Non-parametric analyses, for example those based on ranks or medians, are resistant to the influences of even extreme outliers because error magnitude of ranks and medians is not inflated.

Robust regression detects outliers and provides resistant (stable) results in the presence of outliers by limiting their influence. Three classes of problems have been addressed with robust regression: 1) outliers in the *y*-direction (response direction); 2) outliers in the *x*-direction (covariate space); and, outliers in both directions ^11 pp 8658-8747^.

Common methods for robust regression include M estimation, high breakdown value estimation, and combinations of these two methods ^11 pp 8658-8747^. Huber (1973) introduced M estimation^7^. The method is computationally and theoretically simple. The loss function reduces outliers’ contributions to the squared error loss, thereby limiting their impact on parameter estimates^8^.

Although M estimation is not robust to *x*-direction outliers, it is robust to *y*-direction outliers, and is therefore well-suited to scenarios in which focus is on *y*-direction outliers^7, 8, 12^. Rapid progressors can be considered *y*-direction outliers. Hence, it is not surprising that both the sponsor and FDA implemented robust regression with M estimation as a post-hoc sensitivity analyses of the clinical trials mentioned in Section 1.

## 3. Methods

The objectives of this study are: 1) to characterize the probability of having meaningful imbalance across treatment arms in the number of rapidly progressing patients due to chance alone; 2) assess the influence of imbalances in the number of rapidly progressing patients on estimates of treatment group differences from MMRM; and, 3) to compare results from MMRM with the non-parametric and robust regression methods used in previous AD clinical trials.

### Simulated data

The objectives of this study were pursued via simulation. Two main data scenarios were simulated. First, complete data were simulated and analyzed to assess results without the potentially confounding influence of non-random dropout. A second set of simulations was conducted in data with non-random subject dropout.

The complete data were simulated as a 2 x 3 x 3 factorial arrangement of scenarios, with 2 x 3 = 6 data scenarios, and 3 methods of analysis applied to each data scenario. The simulations included:

- Two levels of magnitude of treatment effect: zero difference between drug and placebo in mean change from baseline and a 0.5pt difference, approximately a 25% slowing of disease progression for drug compared with placebo, which is what was assumed in planning of the aducanumab studies noted in the introduction^1^.
- Three levels (types) of data distribution: a normal distribution and two skewed distributions that were created via a mixture of “normal” and “rapidly progressing” patients; in one skewed distribution the treatment effect was the same in rapid progressors as in the main subgroup, while in the second skewed distribution the treatment effect in rapid progressors was zero.
- Three methods of analysis were used, an MMRM analysis similar to what is commonly used in AD, the Hodges-Lehmann estimator (a non-parametric approach), and robust regression with M estimation.

In each of the data scenarios, 10,000 data sets were simulated with 450 patients per data set, randomized in a 2:1 ratio to the simulated drug and placebo arms, respectively. In each of these data sets no data were missing.

Input parameters for the various simulated data sets are summarized in Tables 1 and 2. Figure 2 is a plot of the Mixed_1 distribution that was a mixture of a main group comprising 95% of the patients with 5% rapid progressors. The distribution in Figure 2 is similar to the distribution of data from the AD clinical trials depicted in Figure 1, indicating that the simulated data provided a reasonable approximation of the clinical trial data.

**Table 1.**
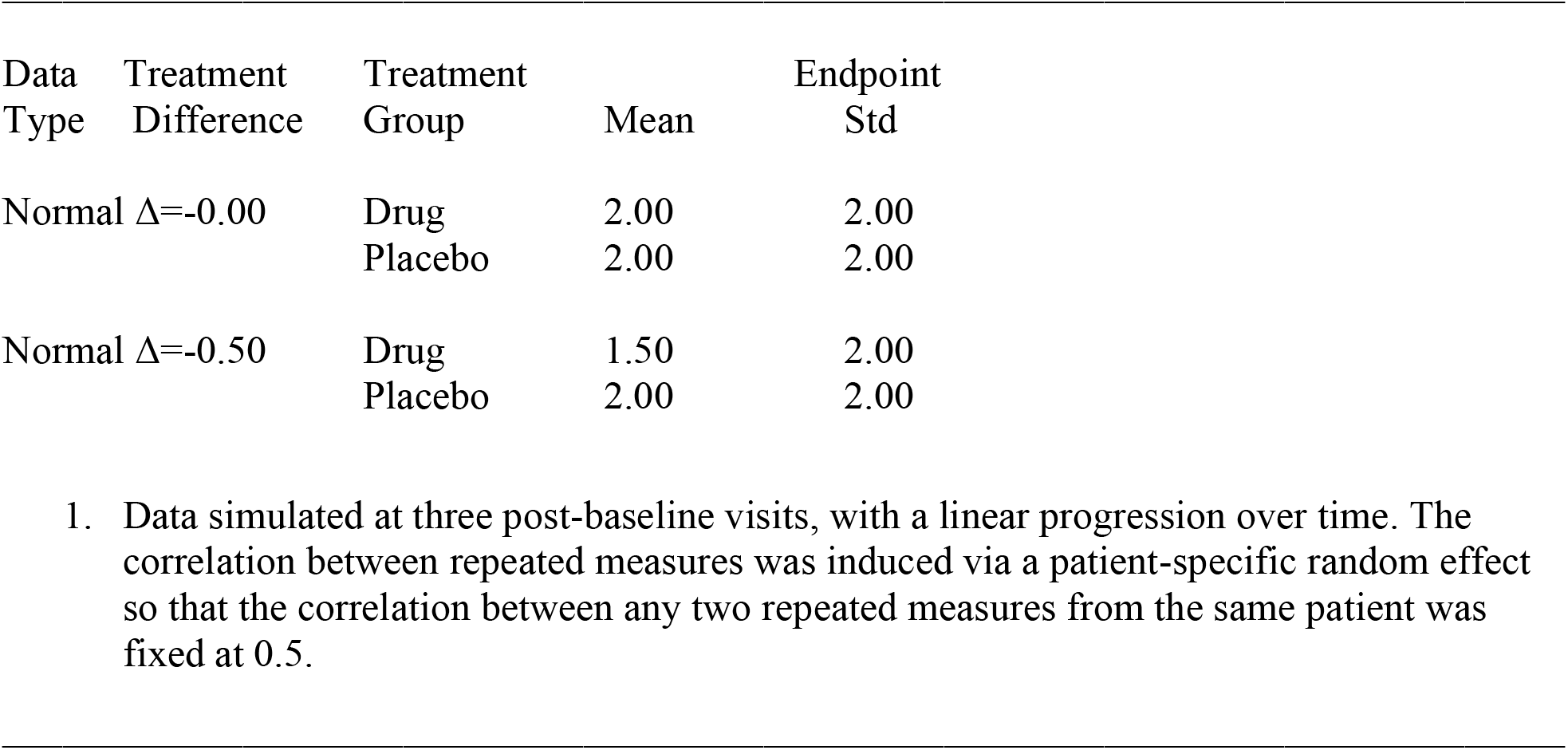
Input parameters for simulations of change from baseline in CDR-sb – normally distributed data^1^.

**Table 2.**
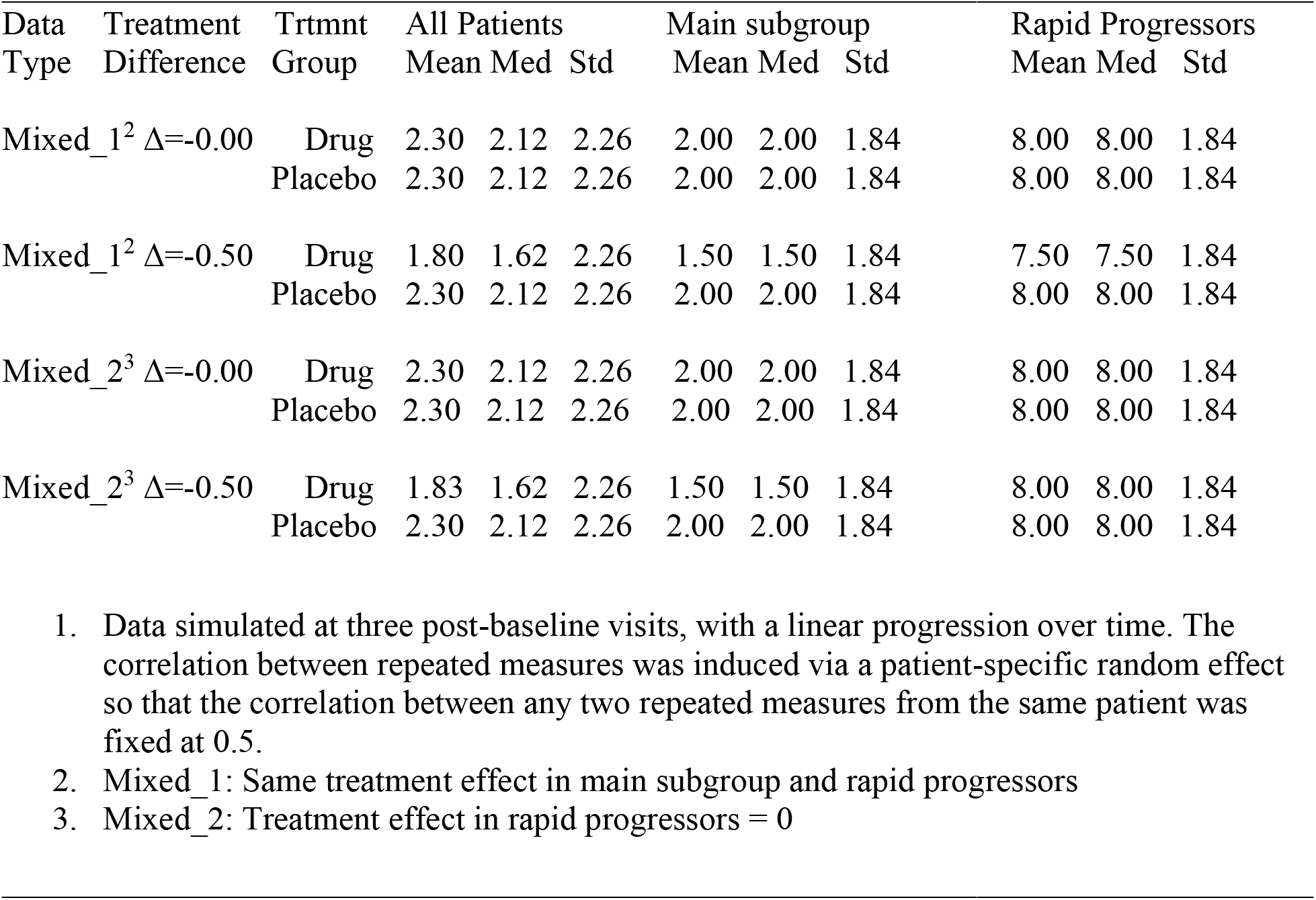
Input parameters for simulations of change from baseline in CDR-sb – Mixed distribution of normal and rapid progressor data^1^.

| Data Type | Treatment Difference | Trtmnt Group | All Patients |  |  | Main subgroup |  |  | Rapid Progressors |  |  |
| --- | --- | --- | --- | --- | --- | --- | --- | --- | --- | --- | --- |
|  |  |  | Mean | Med | Std | Mean | Med | Std | Mean | Med | Std |
| Mixed_1<sup>2</sup> | $\Delta=-0.00$ | Drug | 2.30 | 2.12 | 2.26 | 2.00 | 2.00 | 1.84 | 8.00 | 8.00 | 1.84 |
|  |  | Placebo | 2.30 | 2.12 | 2.26 | 2.00 | 2.00 | 1.84 | 8.00 | 8.00 | 1.84 |
| Mixed_1<sup>2</sup> | $\Delta=-0.50$ | Drug | 1.80 | 1.62 | 2.26 | 1.50 | 1.50 | 1.84 | 7.50 | 7.50 | 1.84 |
|  |  | Placebo | 2.30 | 2.12 | 2.26 | 2.00 | 2.00 | 1.84 | 8.00 | 8.00 | 1.84 |
| Mixed_2<sup>3</sup> | $\Delta=-0.00$ | Drug | 2.30 | 2.12 | 2.26 | 2.00 | 2.00 | 1.84 | 8.00 | 8.00 | 1.84 |
|  |  | Placebo | 2.30 | 2.12 | 2.26 | 2.00 | 2.00 | 1.84 | 8.00 | 8.00 | 1.84 |
| Mixed_2<sup>3</sup> | $\Delta=-0.50$ | Drug | 1.83 | 1.62 | 2.26 | 1.50 | 1.50 | 1.84 | 8.00 | 8.00 | 1.84 |
|  |  | Placebo | 2.30 | 2.12 | 2.26 | 2.00 | 2.00 | 1.84 | 8.00 | 8.00 | 1.84 |

1. Data simulated at three post-baseline visits, with a linear progression over time. The correlation between repeated measures was induced via a patient-specific random effect so that the correlation between any two repeated measures from the same patient was fixed at 0.5. 2. Mixed\_1: Same treatment effect in main subgroup and rapid progressors 3. Mixed\_2: Treatment effect in rapid progressors = 0

**Figure 2.**
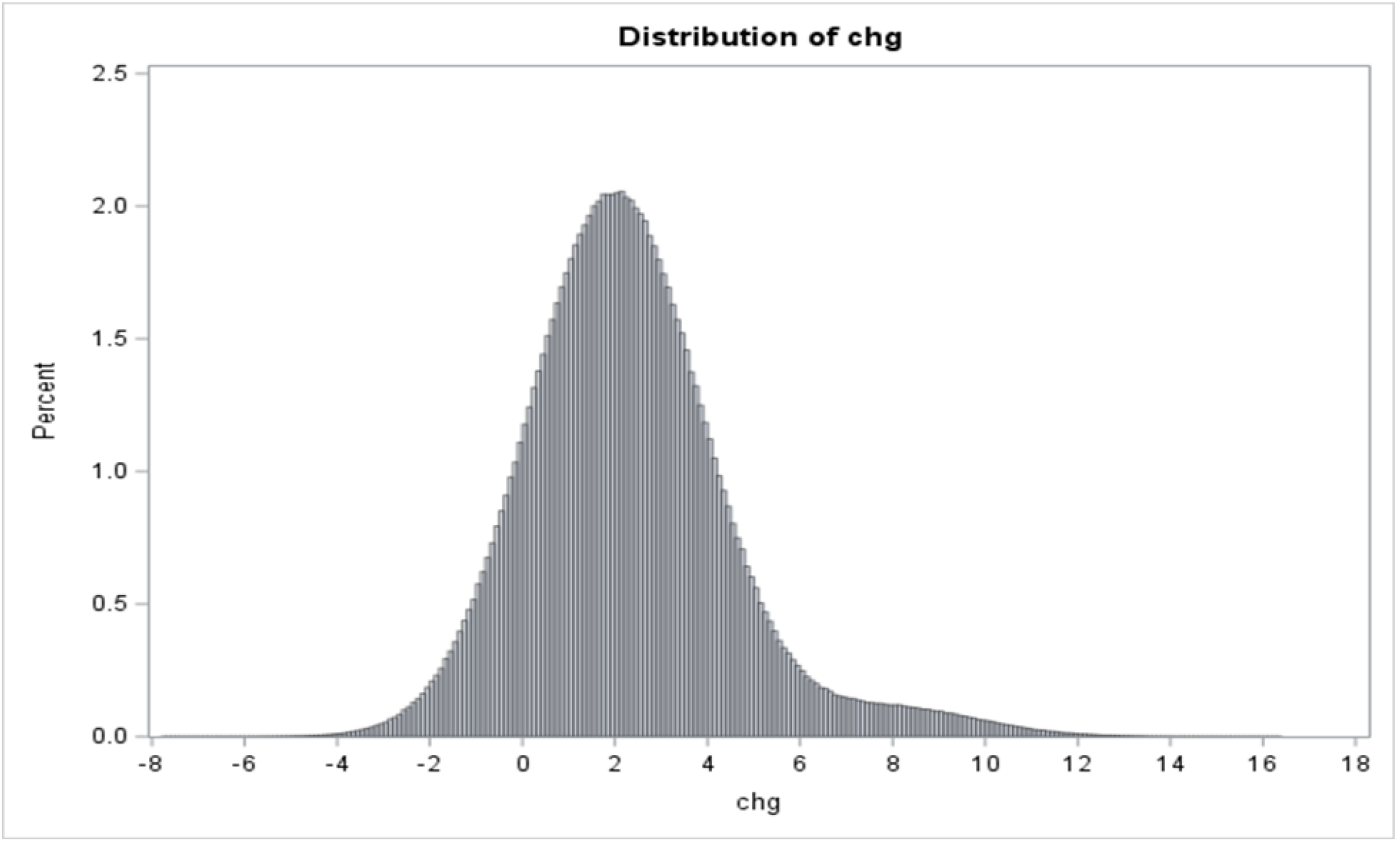
Distribution of simulated outcomes from the Mixed_1 simulations.

An additional set of simulations was conducted to extend results to the more realistic setting of incomplete data. For this simulation the same inputs were used as for the mixed_1 data, except: sample size was 570 and 20% monotone missing data was generated. The mechanism for data deletion was as follows: patients that showed any degree of improvement did not dropout whereas patients that had some degree of worsening had probability of dropout at Visit 2 (and therefore were missing both Visit 2 and Visit 3 observations) of 13%, with an additional 7% drop out at Visit 3 (and therefore were missing only the Visit 3 observation). Because the observations that triggered dropout were deleted, and therefore the observed data did not fully explain the dropout, the missing data mechanism was missing not at random, suggesting some potential for bias in each of the three analytic methods because each assumes a missing at random mechanism. For the missing data scenario, 5000 data sets were simulated rather than 10,000 data sets as in complete data due to the increased computation burden for dealing with missing data in the analyses.

### Analyses

The MMRM analyses were based on SAS PROC MIXED and restricted maximum likelihood estimation^11 pp 6536-6721^. Changes from baseline were modeled using treatment and visit as categorical fixed effects, and baseline score and the baseline score by visit interaction as continuous covariates. Within subject errors were modeled using an unstructured covariance matrix.

The Hodges-Lehmann (HL) approach estimated the median of all pairwise comparisons between all patients in the treated and control groups. As such, it is a non-parametric approach that does not rely on distributional assumptions. The Hodges-Lehmann approach was implemented to test treatment group differences via SAS PROC NPAR1WAY applied to the data at Visit 3 ^11 pp 7122-7194^. Statistical significance was based on whether the 95% confidence interval for the median difference contained 0.

Robust regression (RR) was implemented for the Visit 3 data via PROC ROBUSTREG in SAS using M estimation and the bisquare weighting function ^11 pp 8658-8747^. The model included treatment as a categorical fixed effect and baseline score as a continuous covariate. Estimates were computed using iteratively reweighted least squares (IRLS) with a weighted least squares fit implemented inside an iteration loop. For each iteration, a set of weights for the observations is used in the least squares fit. The weights are constructed by applying the chosen weight function to the current residuals. Initial weights are based on residuals from an initial fit via unweighted least squares. The iteration terminates when a convergence criterion is satisfied ^12^.

In the simulation scenario with missing data, the previously described MMRM analysis was fit to the incomplete data. For RR and HL, an analytic approach similar to that advocated by Mehrotra et al ^6^ was used. Multiple imputation (MI) was implemented via PROC MI ^11 pp 6344-6474^ using 25 rounds of imputation for each of the 5000 simulated data sets, with separate models for each treatment group that included baseline and post-baseline observations. The completed data sets were analyzed using HL and RR as previously described and results were combined using Rubin’s rules as implemented in proc MI analyze^11 pp 6480-6533^.

### Outcomes

Outcomes used to assess results included the mean difference between treatments, the mean standard error of the treatment differences, the standard deviation of the treatment differences, and the percentage of data sets in which the treatment difference was statistically significant (α=0.05). The standard deviation in treatment differences is the empirical standard error of the treatment differences and can be compared with the mean standard error to assess whether the model standard errors accurately reflect the uncertainty in the estimates.

## 4. Results

Results from the normally distributed simulated data are summarized in Table 3. Each method yielded unbiased estimates of treatment group differences when the treatment effect (Δ) was 0.00 and -0.50. The mean standard errors and the standard deviations of the treatment differences were nearly identical within each method, but were greater in HL than in MMRM and RR. The percent of data sets with statistically significant differences was approximately equal to the nominal Type I error rate when Δ = 0.00 for MMRM and RR, and lower than the nominal rate for HL. When Δ = -0.50, the percent of data sets with significant differences (power) was ∼2% greater for MMRM than RR, and 14% greater than HL.

**Table 3.** Results from normally distributed simulated data with no dropout.

|  |  |  |  |  |
| --- | --- | --- | --- | --- |
| $\Delta = 0.00$ | | | | |
| Analysis | Mean $\Delta^1$ | Mean SE<sup>2</sup> | SD $\Delta^3$ | % Significant<sup>4</sup> |
| MMRM | -0.0034 | 0.1737 | 0.1732 | 4.95 |
| HL | -0.0032 | 0.2058 | 0.2057 | 2.38 |
| RR | -0.0034 | 0.1783 | 0.1784 | 4.93 |
| $\Delta = -0.50$ | | | | |
| Analysis | Mean $\Delta^1$ | Mean SE<sup>2</sup> | SD $\Delta^3$ | % Significant<sup>4</sup> |
| MMRM | -0.5034 | 0.1737 | 0.1732 | 82.48 |
| HL | -0.5032 | 0.2058 | 0.2057 | 68.74 |
| RR | -0.5034 | 0.1783 | 0.1784 | 80.45 |

1. Mean estimated treatment difference from the 10,000 datasets 2. Mean standard error of the treatment difference from the 10,000 data sets 3. Standard deviation of the estimated treatment differences from the 10,000 data sets 4. Percent of data sets with a statistically significant treatment difference ( $\alpha=0.05$ )

Results from the mixed_1 set of simulations are summarized in Table 4. Each method yielded unbiased estimates of treatment group differences when Δ was 0.00 and -0.50. The mean standard errors and the standard deviation of the treatment differences were nearly identical within each method, but were lower for RR than for HL and MMRM; that is, in MMRM standard errors were greater than in normally distributed data, but for RR standard errors were similar in the normal and skewed, mixed_1 simulated data sets. With Δ = 0.00, trends for Type I error were similar to those in normally distributed data. When Δ = -0.50, unlike in normally distributed data where power was similar for MMRM and RR, power was ∼12% greater for RR than for MMRM, with MMRM being similar to HL. Although the average Δ was similar across methods, in 20% of the data sets the estimate from MMRM differed from the corresponding estimate in RR and HL by at least 0.15. In other words, although average estimates were similar, it was not unusual for the various analyses to have meaningful differences within datasets.

**Table 4.** Results from the mixed_1 simulated data with no dropout.

| $\Delta = 0.00$ | | | | | |
| --- | --- | --- | --- | --- | --- |
| Analysis | Mean $\Delta^1$ | Mean SE<sup>2</sup> | SD $\Delta^3$ | % Significant<sup>4</sup> | 80% Interval<sup>5</sup> |
| MMRM | -0.0004 | 0.2067 | 0.2088 | 5.28 |  |
| HL | -0.0004 | 0.2083 | 0.2082 | 2.39 | $\pm 0.15$ |
| RR | -0.0004 | 0.1787 | 0.1805 | 5.32 | $\pm 0.15$ |
| $\Delta = -0.50$ | | | | | |
| Analysis | Mean $\Delta^1$ | Mean SE<sup>2</sup> | SD $\Delta^3$ | % Significant<sup>4</sup> | 80% Interval<sup>5</sup> |
| MMRM | -0.5004 | 0.2067 | 0.2088 | 67.75 |  |
| HL | -0.5034 | 0.2083 | 0.2082 | 67.34 | $\pm 0.15$ |
| RR | -0.5004 | 0.1787 | 0.1805 | 79.47 | $\pm 0.15$ |
| 1. Mean estimated treatment difference from the 10,000 datasets<br>2. Mean standard error of the treatment difference from the 10,000 data sets<br>3. Standard deviation of the estimated treatment differences from the 10,000 data sets<br>4. Percent of data sets with a statistically significant treatment difference ($\alpha=0.05$)<br>5. For each data set, the estimated treatment contrast was compared between MMRM and the other methods. The 80% interval defines the range that included 80% of the pairwise comparisons within data sets. |  |  |  |  |  |

Results from the mixed_1 set of simulations are further summarized by sub-setting the datasets according to the ratio of rapid progressing patients in the two treatment arms. Table 5 summarizes results when Δ = 0.00 and Table 6 summarizes results when Δ = -0.50.

**Table 5.**
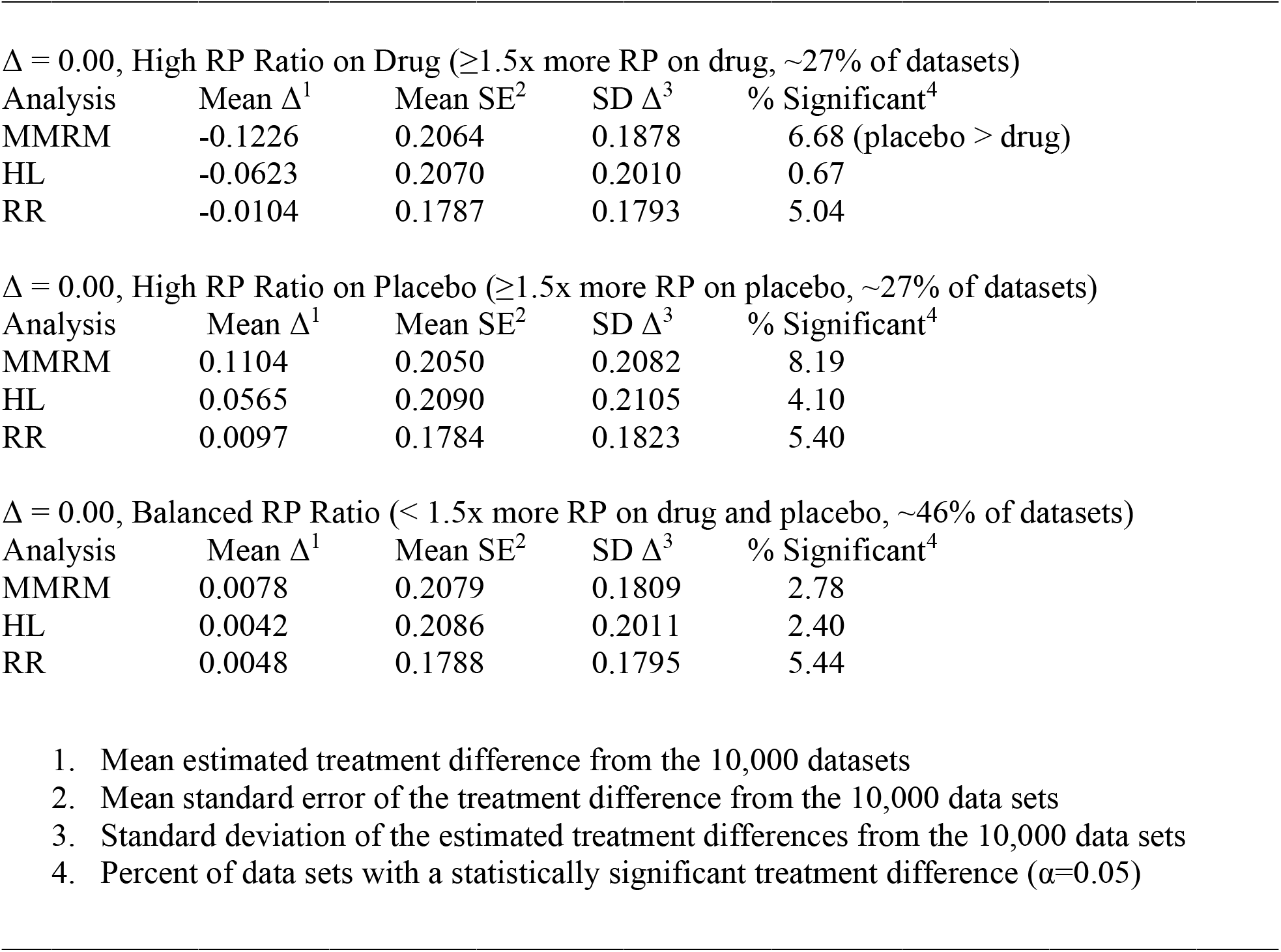
Results from the mixed_1 simulated data with no dropout and Δ = 0.00, by data subgroups defined by the ratio of rapidly progressing patients in the drug and placebo arms.

| Analysis | Mean $\Delta^1$ | Mean SE<sup>2</sup> | SD $\Delta^3$ | % Significant<sup>4</sup> |
| --- | --- | --- | --- | --- |
| MMRM | -0.1226 | 0.2064 | 0.1878 | 6.68 (placebo > drug) |
| HL | -0.0623 | 0.2070 | 0.2010 | 0.67 |
| RR | -0.0104 | 0.1787 | 0.1793 | 5.04 |

$\Delta = 0.00$ , High RP Ratio on Placebo ( $\geq 1.5x$ more RP on placebo, ~27% of datasets)
| Analysis | Mean $\Delta^1$ | Mean SE<sup>2</sup> | SD $\Delta^3$ | % Significant<sup>4</sup> |
| --- | --- | --- | --- | --- |
| MMRM | 0.1104 | 0.2050 | 0.2082 | 8.19 |
| HL | 0.0565 | 0.2090 | 0.2105 | 4.10 |
| RR | 0.0097 | 0.1784 | 0.1823 | 5.40 |

$\Delta = 0.00$ , Balanced RP Ratio ( $< 1.5x$ more RP on drug and placebo, ~46% of datasets)
| Analysis | Mean $\Delta^1$ | Mean SE<sup>2</sup> | SD $\Delta^3$ | % Significant<sup>4</sup> |
| --- | --- | --- | --- | --- |
| MMRM | 0.0078 | 0.2079 | 0.1809 | 2.78 |
| HL | 0.0042 | 0.2086 | 0.2011 | 2.40 |
| RR | 0.0048 | 0.1788 | 0.1795 | 5.44 |

1. Mean estimated treatment difference from the 10,000 datasets 2. Mean standard error of the treatment difference from the 10,000 data sets 3. Standard deviation of the estimated treatment differences from the 10,000 data sets 4. Percent of data sets with a statistically significant treatment difference ( $\alpha=0.05$ ) -

**Table 6.**
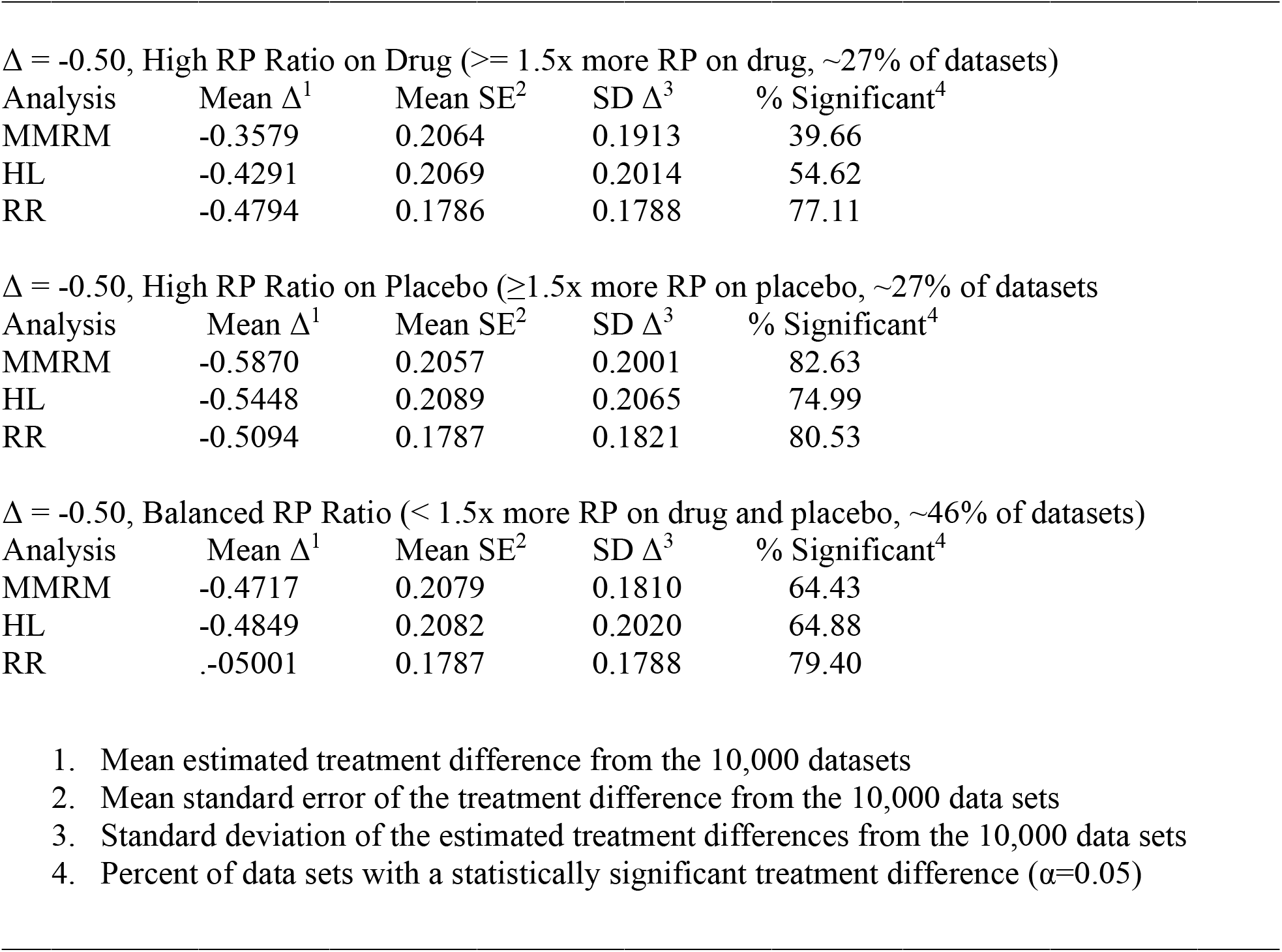
Results from the mixed_1 simulated data with no dropout and Δ = -0.50, by data subgroups defined by the ratio of rapidly progressing patients in the drug and placebo arms.

The mean Δs from MMRM varied consistent with the ratio or rapid progressing patients. In the subset of datasets in which the RP ratio was ≥ 1.5x on drug (more rapid progressors in the drug arm) the mean Δ from MMRM was less than the corresponding simulation input values of Δ = 0.00 (Table 6) and Δ = -0.50 (Table 7). The opposite trend existed when the RP ratio was ≥ 1.5x on placebo (more rapid progressors in the placebo arm), with the mean Δ from MMRM being greater than the corresponding simulation input values. In contrast, mean Δ’s from RR did not appreciably vary across the categories of data subsets defined by RP ratio. Results for HL were intermediate to those of MMRM and RR. When the ratio of RPs did not appreciably differ mean Δ’s from each method of analysis was close to the simulation input values.

**Table 7.** Results from the mixed_1 simulated data with 20% dropout.

|  |  |  |  |  |
| --- | --- | --- | --- | --- |
| $\Delta = 0.00$ | | | | |
| Analysis | Mean $\Delta^1$ | Mean SE<sup>2</sup> | SD $\Delta^3$ | % Significant<sup>4</sup> |
| MMRM | -0.0032 | 0.2003 | 0.2006 | 5.00 |
| HL | -0.0033 | 0.2092 | 0.2042 | 4.38 |
| RR | -0.0034 | 0.1858 | 0.1807 | 4.30 |
| $\Delta = -0.50$ | | | | |
| Analysis | Mean $\Delta^1$ | Mean SE<sup>2</sup> | SD $\Delta^3$ | % Significant<sup>4</sup> |
| MMRM | -0.5166 | 0.1996 | 0.2004 | 73.56 |
| HL | -0.5199 | 0.2087 | 0.2042 | 70.74 |
| RR | -0.5178 | 0.1848 | 0.1508 | 80.42 |

1. Mean estimated treatment difference from the 5,000 datasets 2. Mean standard error of the treatment difference from the 5,000 data sets 3. Standard deviation of the estimated treatment differences from the 5,000 data sets 4. Percent of data sets with a statistically significant treatment difference ( $\alpha=0.05$ ) -

Given the way data were subset based on RP ratio, mean Δs differing from simulation input values is not a valid measure of bias, but rather a means of assessing stability (consistency) of results.

Results from the mixed_2 set of simulations (no treatment effect in the rapidly progressing patients) are summarized in Table S2 by grouping the datasets according to the ratio of rapid progressing patients in the two treatment arms. Results followed the same pattern as in the mixed_1 set of simulations with mean Δs from MMRM varying consistent with the ratio of rapid progressing patients whereas results from RR were consistent across data set groupings. See the supplemental material for more details.

Results from the mixed_1 set of simulations in data with 20% dropout are summarized in Table 7. Results followed the same pattern as in the mixed_1 and mixed_2 sets of simulations that had no dropout. The advantage of RR over MMRM and HL in power was ∼7% and ∼10%, respectively. Each method provided control of Type I error at the nominal rate (or slightly less) The average estimated treatment contrast was slightly greater than the input value when the treatment effect was -0.50 because the analyses assumed a missing at random mechanism when the actual mechanism was missing not at random.

## 5. Discussion

Statistical theory suggests methods other than MMRM may be useful to consider when data are heavily skewed ^6^. as they were in the AD clinical trials that motivated this investigation Via the central limit theorem, the concern in large trials regarding non-normal data is not bias, the concern is stability of results ^7, 8, 9, 10^, although bias could be a concern in small trials.

This investigation showed that 1) chance alone can often result in substantial imbalance across treatment arms in the number of rapid progressing patients; and, 2) these imbalances can influence estimates of treatment group differences. In over half the simulated data sets with complete data (and hence no confounding from non-random dropout) the ratio of rapid progressing patients was at least 1.5-fold greater on one arm than the other; and, treatment group differences estimated via MMRM varied in accordance with the ratio of rapid progressors on drug versus placebo. Therefore, the imbalance and its consequences seen in the recent AD clinical trials should be anticipated in planning AD studies.

This study also provided evidence, consistent with statistical theory, supporting the usefulness of robust methods. In normally distributed data, MMRM had ∼2% more power than RR and ∼14% more power than HL. As expected, MMRM and RR each controlled Type I error at the nominal level, with HL having ∼1/2 the nominal rate.

In the skewed data, mimicking the AD clinical trials, each of the methods yielded unbiased estimates of the treatment contrast, but standard errors were smaller and power was greater for RR than for MMRM and HL. When the data sets were subset by the ratio of rapid progressing patients across treatment arms, the average results from MMRM varied in accordance with this ratio. The average treatment effect in MMRM was greater when the ratio of rapid progressors was higher on placebo, and the average treatment effect was lower with more rapid progressors on drug. Results from HL showed the same trend, but to a lesser degree than MMRM. Robust regression yielded the most stable results, with smaller average standard errors and similar average treatment contrasts regardless of the ratio of rapid progressors. Similar results were seen in the scenario with 20% missing data.

These results should be interpreted considering several limitations. Although the simulated data were similar to the clinical trial data that motivated this investigation, information on the distribution of outcomes from other AD clinical trials is lacking. It may be that a log-normal or Cauchy distribution better describes AD data than the mixture distribution used here to simulate data. Moreover, other analytic approaches should be considered for assessing mean changes. For example, mixture models might perform better than RR if a mixture distribution best describes AD data. Or, if the data are best described by a log-normal distribution, a log-transformation prior to an MMRM analysis may be better. Or, perhaps quantile regression is a useful alternative.

Therefore, further investigation is needed to compare the strengths and limitations of analytic options over a wider set of conditions. However, results of this investigation suggest that imbalances across treatment arms in the number of rapid progressors is likely in AD clinical trials. These imbalances influence results from MMRM, and it should not be assumed that MMRM is the optimum or only analysis needed in AD clinical trials.

## Data Availability

All data produced in the present study are available upon reasonable request to the authors

## Acknowledgements

None

## Funding

No funding sources to declare

## Declaration of interests statement

There are no competing interests to declare.

## Appendix - Supplementary Material

**Table S1.** Results from the primary and secondary endpoints in the aducanumab phase 3 clinical trials.

| Endpoint | EMERGE |  |  | ENGAGE |  |  |
| --- | --- | --- | --- | --- | --- | --- |
|  | Placebo<br>decline ± SE<br>(n=548) | Difference vs placebo (%)<sup>a</sup><br>95% CI<br>*P* value |  | Placebo<br>decline ± SE<br>(n=545) | Difference vs placebo (%)<sup>a</sup><br>95% CI<br>*P* value |  |
|  |  | Low dose<br>(n=543) | High dose<br>(n=547) |  | Low dose<br>(n=547) | High dose<br>(n=555) |
| Primary |  |  |  |  |  |  |
| CDR-SB<sup>a</sup> | 1.74±0.11 | −0.26 (−15)<br>−0.57, 0.04<br>.090 | −0.39 (−22)<br>−0.69, −0.09<br>.012 | 1.56±0.11 | −0.18 (−12)<br>−0.47, 0.11<br>.225 | 0.03 (2)<br>−0.26, 0.33<br>.833 |
| Secondary |  |  |  |  |  |  |
| MMSE<sup>b</sup> | −3.3±0.2 | −0.1 (3)<br>−0.7, 0.5<br>.758 | 0.6 (−18)<br>0.0, 1.1<br>.049 | −3.5±0.2 | 0.2 (−6)<br>−0.3, 0.7<br>.479 | −0.1 (3)<br>−0.6, 0.5<br>.811 |
| ADAS-Cog13<sup>c</sup> | 5.16±0.40 | −0.70 (−14)<br>−1.76, 0.36<br>.196 | −1.40 (−27)<br>−2.46, −0.34<br>.010 | 5.14±0.38 | −0.58 (−11)<br>−1.58, 0.42<br>.254 | −0.59 (−11)<br>−1.61, 0.43<br>.258 |
| ADCS-ADL-MCI<sup>d</sup> | −4.3±0.4 | 0.7 (−16)<br>−0.3, 1.7<br>.151 | 1.7 (−40)<br>0.7, 2.7<br><.001 | −3.8±0.3 | 0.7 (−18)<br>−0.2, 1.6<br>.123 | 0.7 (−18)<br>−0.2, 1.6<br>.151 |

**Table S2.** Results from the mixed_2 simulated data with Δ = -0.50 by groups defined by the ratio of rapidly progressing patients in the drug and placebo arms.

| Analysis | Mean $\Delta^1$ | Mean SE<sup>2</sup> | SD $\Delta^3$ | % Significant |
| --- | --- | --- | --- | --- |
| MMRM | -0.3515 | 0.2121 | 0.1868 | 36.94<sup>4</sup> |
| HL | -0.4347 | 0.2075 | 0.2001 | 55.06 |
| RR | -0.5009 | 0.1775 | 0.1772 | 80.27 |

$\Delta = -0.50$ , High RP Ratio on Placebo ( $\geq 1.5x$ more RP on placebo, 27% of datasets)
| Analysis | Mean $\Delta^1$ | Mean SE<sup>2</sup> | SD $\Delta^3$ | % Significant<sup>4</sup> |
| --- | --- | --- | --- | --- |
| MMRM | -0.5901 | 0.2091 | 0.2059 | 81.64 |
| HL | -0.5565 | 0.2090 | 0.2061 | 75.64 |
| RR | -0.5293 | 0.1774 | 0.1814 | 84.13 |

$\Delta = -0.50$ , Balanced RP Ratio ( $< 1.5x$ more RP on drug and placebo, 46% of datasets)
| Analysis | Mean $\Delta^1$ | Mean SE<sup>2</sup> | SD $\Delta^3$ | % Significant<sup>4</sup> |
| --- | --- | --- | --- | --- |
| MMRM | -0.4785 | 0.2127 | 0.1834 | 62.51 |
| HL | -0.5017 | 0.2089 | 0.2020 | 67.41 |
| RR | -0.5204 | 0.1780 | 0.1786 | 83.02 |

1. Mean estimated treatment difference from the 10,000 datasets 2. Mean standard error of the treatment difference from the 10,000 data sets 3. Standard deviation of the estimated treatment differences from the 10,000 data sets 4. Percent of data sets with a statistically significant treatment difference ( $\alpha=0.05$ ) -

## Notes

### Competing Interest Statement

The authors have declared no competing interest.

### Funding Statement

This study did not receive any funding

